# Using remotely delivered Spring Forest Qigong™ to reduce neuropathic pain in adults with spinal cord injury: A non-randomized controlled trial

**DOI:** 10.1101/2023.02.11.23285793

**Authors:** Ann Van de Winckel, Sydney T. Carpentier, Wei Deng, Lin Zhang, Angela Philippus, Kimberley R. Monden, Ricardo Battaglino, Leslie R. Morse

## Abstract

**Importance:** The manuscript proposes the feasibility and potential of a remote Qigong intervention to reduce neuropathic pain in adults with spinal cord injury (SCI)-related neuropathic pain.

**Objective:** We determined the feasibility and estimates of efficacy of a remotely delivered Qigong intervention in adults with SCI-related neuropathic pain.

**Design:** This is a non-randomized controlled trial with outcomes assessed at baseline-, 6- and 12-weeks of Qigong practice, and at 6-weeks and 1-year follow-up.

**Setting:** Completely remote clinical trial.

**Participants:** Adults with SCI-related neuropathic pain, with SCI ≥3 months, with complete or incomplete SCI, and highest neuropathic pain level of >3 on the Numeric Pain Rating Scale (NPRS). We used nationwide volunteer sampling.

We recruited 23 adults with chronic SCI (7/2021-2/2022). Eighteen participants started the study and completed all study components, including the 6-week follow-up. Twelve participants completed the 1-year follow-up assessment.

**Intervention:** Participants practiced the Spring Forest Qigong™ “Five Element Healing Movements” with an online video by combining movement with kinesthetic imagery, at least 3x/week for 12 weeks.

**Main Outcome(s) and Measure(s):** To address the feasibility outcome and track adherence, the website automatically monitored the days and duration that the Qigong video was played. Self-report neuropathic pain intensity and SCI-related symptoms such as spasms, functional performance, mood, and body appreciation were also collected.

**Results:** Eighteen participants, 60±12 years of age, 15±11 years post-SCI had a highest baseline *neuropathic pain* of 7.94±2.33 on the NPRS, which was reduced to 4.17±3.07 after 12 weeks of Qigong practice (Cohen’s *d*=1.75). This pain relief remained at 6-week and 1-year follow-ups. Participants reported reduced spasm frequency (change score 1.17±1.20, *d*=0.98) and severity (0.72±1.02, *d*=0.71), and reduced interference of neuropathic pain on mood (3.44±2.53, *d*=1.36), sleep (3.39±2.40, *d*=1.41), and daily activities (3.17±2.77, *d*=1.14). They had a greater ability to perform functional activities (Patient Specific Functional Scale, 6.68±3.07, *d*=2.18) and had improved mood (Patient Health Questionnaire-9, 2.33±3.31, *d*=0.70).

**Conclusions and Relevance:** Our preliminary data demonstrate the feasibility of Qigong practice in adults with SCI-related neuropathic pain and promising results of neuropathic pain relief and improvement in SCI-related symptoms after Qigong practice.

**Trial Registration (this manuscript refers to the quasi-experimental substudy):** CREATION: A Clinical Trial of Qigong for Neuropathic Pain Relief in Adults with Spinal Cord Injury, NCT04917107, https://www.clinicaltrials.gov/ct2/show/NCT04917107.

## 1. INTRODUCTION

There are about 299,000 adults with spinal cord injury (SCI) in the United States (US).^1^ Pain is one of the most common secondary health consequences of SCI and the most difficult to treat.^2^ This is especially true for chronic neuropathic pain, which is described as sharp, shooting, stabbing, electric, or burning, and sometimes excruciating pain at, below, or above the level of spinal injury.^2,3^ The prevalence of neuropathic pain in adults with SCI is high: up to 69% of adults with SCI deal with debilitating chronic neuropathic pain, which interferes with rehabilitation and quality of life.^4–8^ Accessible treatment options such as exercise and medications have limited success in reducing pain. For instance, pain medications result in less than 50% reduction in pain for only about one-third of the people using them.^2,9,10^ Studies with SCI stakeholders confirm that accessible treatments are limited and primarily focused on pain medications with insufficient benefit but with significant risks for addiction and adverse effects.^11,12^ Stakeholders further reported that improved patient access to non-pharmacological treatment options is urgently needed.^11,12^

While the exact mechanisms of neuropathic pain are unknown, it is believed that changes in brain function may contribute to neuropathic pain.^13–17^ Concurrently with reduced or absent sensation, recently, increasing evidence shows that adults with SCI experience deficits in body awareness.^18–31^ Body awareness deficits are thought to contribute to the production or maintenance of chronic neuropathic pain.^18–20,22,24,25^ Body awareness refers to an attentional focus on and awareness of internal body sensations, which includes an awareness of how the body and body parts are positioned and move in space.^21,32^

Mind and body approaches are known to improve body awareness and thus could be a viable approach to treating neuropathic pain.^32–37^ However, research demonstrating the effectiveness of mind and body approaches for reducing pain in SCI is limited to seven studies among which only three (Yoga and Tai Chi)^38–40^ looked at reducing pain but without mentioning which type of pain they were investigating.^38–44^ Mind and body research in SCI is fairly recent, and often movement modifications are needed to allow participation by adults with SCI. Chalageri *et al*. (2021) showed significant pain reduction after Yoga meditation with conventional rehabilitation, with the most benefit for those with acute SCI and paraplegia.^38^ Curtis *et al*. (2017) reported greater improvements in depressive symptoms and self-compassion, but not in pain, in adults with SCI doing adaptive yoga vs. those in a waitlist group.^40^ Shem *et al*. (2016) reported that a seated Tai Chi program was well-tolerated in adults with SCI with benefits in pain, and emotional and physical sense of well-being after each session.^39^ However, the weekly in-person classes were associated with a drop-out rate of 60%. Tsang *et al*. (2015) reported greater improved dynamic sitting balance performance and handgrip strength after 12 weeks of sitting Tai Chi training in 11 adults with SCI compared to 8 controls.^42^ With the exception of Chalageri *et al*.’s study with Yoga (n=91), the sample sizes in these studies were small (n=23, n=26, n=19), and only Tsang *et al*. (2015)’s study included adults with incomplete tetraplegia; all others recruited adults with paraplegia only. These are promising results, however, more and larger studies with accessible interventions for adults with tetraplegia are needed to determine the potential efficacy of these interventions to alleviate neuropathic pain.

Of the mind and body approaches, Qigong seems to be the most accessible approach for adults with SCI due to the simple, gentle movements, combined with a focus on breathing and body awareness. Remote delivery of Qigong provides a feasible approach for adults with SCI as it eliminates the commonly reported barriers to participating in in-person interventions: fatigue, pain, transportation, and scheduling difficulties.^45^

Therefore, our first objective of this study was to determine the feasibility of a 12-week remotely delivered Spring Forest Qigong™ “Five Element Healing Movements” in adults with SCI-related neuropathic pain. This Qigong program can be practiced in standing, sitting, or lying positions.^46,47^ Moreover, our remote Qigong practice at home with an online video combines actively moving to the participant’s ability level with kinesthetic imagery, i.e., focusing on the feeling of moving the whole body *<u>as if</u>* in an upright position, because imagery may be an additional way to improve body awareness and reduce pain.^22,23^ Therefore, adults with high-level tetraplegia after SCI can participate in this study because they can easily practice Qigong in their power wheelchair or lying down. In contrast, in other studies, the requirement of minimum active muscle strength to perform the exercises often excludes these individuals. Our second objective was to calculate estimates of efficacy of this remotely delivered Qigong intervention in adults with SCI-related neuropathic pain to inform future efficacy clinical trials.

## 2. METHODS

The details of the protocol can be found in Van de Winckel *et al*. (2022).^48^

### 2.1 Design

We performed a non-randomized controlled trial (remote-only substudy of the CREATION trial) in which participants were assessed at baseline for neuropathic pain and SCI-related symptoms, after 6- and 12-weeks of Qigong practice at home, at least 3x/week for 41min/session, and with follow-up assessments at 6-weeks and 1-year post-Qigong practice.

### 2.2 Setting

This is an entirely remote clinical trial. Participants are community-dwelling adults with SCI and neuropathic pain. Participants practiced at home or in any location of their choice that had an internet connection.

### 2.3 Recruitment

We used volunteer sampling nationwide through fliers and announcements on relevant websites, as well as letters sent through the M Health/Fairview healthcare system in the Twin Cities, MN. Participants were also recruited in outpatient locations within a newly funded Spinal Cord Injury Model System Center (Minnesota Regional Spinal Cord Injury Model System) and the community. Specifically, we have collaborators within the Minneapolis VA Health Care System including the Minnesota Paralyzed Veterans; M Health/Fairview (Minneapolis); MN SCI associations; Regions Hospital; Courage Kenny Rehabilitation Institute; Get Up Stand Up To Cure Paralysis; Unite2Fight paralysis, and Fit4Recovery.

### 2.4 Participants

We recruited adults with SCI-related neuropathic pain, with SCI of more than 3 months, who were medically stable with complete or incomplete SCI, and with the highest level of SCI-related neuropathic pain in the prior week, higher than 3 out of 10 on the Numeric Pain Rating Scale.^49^

### 2.5 Consenting, ethical approvals, screening, and baseline characteristics of participants

After a phone screening, eligible participants had a virtual visit through the UMN secure Zoom conference platform with the study staff to review and address questions about the combined HIPAA/informed eConsent. The eConsent was signed through a UMN Research Electronic Data Capture (REDCap) link. REDCap uses a MySQL database via a secure web interface with data checks to ensure data quality during data entry.

The study was conducted in accordance with the principles of the Declaration of Helsinki (2013)^50^ and the Institutional Review Board (IRB) of the University of Minnesota approved the study (IRB# STUDY00011997). The CONSORT reporting guidelines were followed.^51,52^

The study staff member acquired demographic information and general health and medical history from the participants. The study staff screened for cognitive impairments with the Mini-Mental State Examination-short version (cut-off score <13/16).^53,54^ We also tested their ability to perform kinesthetic motor imagery with the Kinesthetic and Visual Imagery Questionnaire (cut-off score <15/25 points).^55^ Data was collected on REDCap.

### 2.6 Intervention

The Spring Forest Qigong™ “Five Element Healing Movements” video, https://www.springforestqigong.com/, was developed by Qigong Grand Master Chunyi Lin, MS in holistic healing, who founded the Spring Forest Qigong™ center in Minnesota in 1995 after decades of study with some of the most renowned Qigong masters in China. Grand Master Lin demonstrates in the video gentle horizontal and vertical arm and leg movements in specific postures in standing position. The Qigong movements are known to strengthen the legs and core muscles, and improve body awareness, balance, and posture while relaxing the body with a combined focus on breathing and movement.^46,56^ The movements also help restore emotional balance.^46,56,57^

A Qigong Master from the Spring Forest Qigong™ Center (Minnesota) taught the introductory class of the “Five Element Healing Movements” (6 hours) over Zoom. Then participants logged in to the Spring Forest Qigong™ website with their unique study identification number and password to access the Spring Forest Qigong™ “Five Element Healing Movements” video and were asked to practice at least 3x/week for 41min/session. The five Qigong movements are Movement 1: Moving of Yin and Yang; Movement 2: Breathing of the Universe; Movement 3: Connecting with Heaven & Earth; Movement 4: Connecting with Your Body’s Energy; Movement 5: Connecting with Your Heart’s Energy. Details of how the different movements are performed are presented in our protocol paper.^48^

The movements were accessible to adults with SCI with paraplegia or tetraplegia because the Qigong movements can also be done lying or sitting. Additionally, the first author (AVDW) developed specific kinesthetic imagery instructions to allow participants with all levels of mobility to participate maximally in the Qigong practice. The participants were instructed to actively move along with the video however much they comfortably could and to perform kinesthetic imagery at the same time. The instructions of the kinesthetic imagery were to focus on the feeling of the body as if they were standing up and doing the whole-body movement. For example, they were asked to imagine the ‘feeling’ of the contact of the floor with the soles of their feet and the feeling of the flow of the movements in the whole body, rather than to ‘visualize’ the whole-body movement. If reproducing this feeling was difficult, participants were asked to associate positive memories from before their SCI with this imagined standing posture. For example, positive memories of feeling the warm sand under the soles of their feet when walking on the beach or the feeling of grass under the soles of bare feet when playing in the garden with children or grandchildren.

### 2.7 Follow-up period

During the 6-week follow-up, we asked the participants to stop practicing Qigong, but they could continue taking pain medication if they needed any, and they received regular standard of care as needed. After the 6-week follow-up period, participants could restart their Qigong practice at the frequency of their choice. We did a final assessment of neuropathic pain and function at 1-year follow-up to evaluate any change in behavior occurring when no instructions were given. Participants received $100 upon study completion (at the 6-week follow-up), through a Greenphire ClinCard, and an additional $25 if they also completed the 1-year follow-up. The Qigong introduction class and video access were paid for by the study.

### 2.8 Main Outcome(s) and Measure(s)

Based on models and guidelines for intervention development,^58,59^ and on prior publications related to ***feasibility markers***,^60–63^ we assessed the following *a-priori* feasibility indicators: We estimated recruitment of 40% of adults with complete SCI and 60% of adults with incomplete SCI; Qigong adherence of at least 70% of participants practicing at least 2x/week; maximum 30% attrition (given that the study started in the midst of the COVID-19 pandemic); none of the questionnaires fully missing in more than 25% of the participants; mild adverse events related to the study in maximum 10% of the participants; and 70% or more participants satisfied with the program. Quotes from participants were used to elucidate our findings on satisfaction with the Qigong practice and the program.

To ***track adherence*** to the intervention, the Spring Forest Qigong™ website automatically monitored the days and duration that videos were accessed. Logs with the participant’s codes and days/times practiced were provided to the investigators. Additionally, the first author (AVDW) did weekly check-ins with the participants to address any questions about the Qigong practice and to demonstrate movements over Zoom if needed. Participants were asked to report any adverse events and potential perceptions, effects, and satisfaction with the Qigong practice. The first author (AVDW) is level 5 (out of 5) in the Spring Forest Qigong™ curriculum and is a certified practice group leader and has practiced Qigong for about 10 years.

Given the slow, gentle movements and kinesthetic imagery, and weekly check-ins, the ***risks*** of Qigong practice were considered to be minimal and expected to be limited to mild transient discomfort. A plan was in place to address any (serious) adverse events as per IRB and Good Clinical Practice requirements. If participants were prescribed neuropathic pain medication prior to the start of the study they were permitted to continue taking those. However, in order to avoid bias from concomitant interventions, other health appointments for neuropathic pain (e.g., osteopathy) were not scheduled or permitted during the duration of the study.

***As the primary outcome measure***, graduate students called the participants weekly to ask about their highest, average, and lowest *neuropathic pain* intensity ratings perceived in the prior week, using the 0-10 Numerical Pain Rating Scale with 0 being no pain and 10 being the worst pain ever.^49^ This scale has been recommended by the mission of the Initiative on Methods, Measurement, and Pain Assessment in Clinical Trials consensus group for use in pain clinical trials and by the 2006 National Institute on Disability and Rehabilitation Research SCI Pain outcome measures consensus group.^64,65^ Distinctions between neuropathic pain, nociceptive musculoskeletal pain, or nociceptive other pain (e.g., visceral pain) were made using the National Institute of Neurological Disorders-Common Data Elements International SCI Pain Basic Data Set Version 2.0.^66^ During the weekly call, we also assessed the dosage of neuropathic pain medication taken that week, recent illnesses, health care utilization, and/or recent hospitalizations.

The following ***secondary outcome measures*** were collected by graduate students over Zoom at five time points: baseline, 6-weeks (mid-Qigong practice), 12-weeks (end of Qigong practice), 6-week follow-up, and 1-year follow-up.

The National Institute of Neurological Disorders-Common Data Elements International SCI Pain Basic Data Set Version 2.0^66^ also assessed dimensions of pain (i.e., the intensity of pain, location, etc.), and how pain interferes with mood, activity, and sleep.

Frequency of spasms and spasm severity were evaluated with the Penn Spasm Frequency Scale.^67^ Mood (i.e., symptoms of depression) was assessed with the Patient Health Questionnaire-9 (PHQ-9).^68–72^ Body appreciation was assessed with the Functional Appreciation Scale (FAS), which measures the level of appreciation one has for the functionality of the body.^73^

Participants provided a self-report of whether general cardiovascular function, bladder, bowel, and sexual function were normal, abnormal, or absent with the Autonomic Standards Assessment Form.^74^

Function was assessed with the Spinal Cord Injury Functional Index,^75^ measuring functional performance across 5 domains: basic mobility, self-care, fine motor function, and ambulation.^76^ This scale has separate questions for adults with paraplegia and tetraplegia. For the Patient Specific Functional Scale (PSFS),^77^ the participants self-identified goals related to activities in daily life that were important to them but that they had difficulty doing because of the neuropathic pain. The participants rated them at each time point between 0 (unable to do the activity without pain) and 10 (able to do the activity without pain).^77^

### 2.9 Sample size

To our knowledge, this is the first study evaluating the feasibility of Qigong on neuropathic pain perception in adults with SCI. This study also provides preliminary estimates of effect size needed to generate hypotheses for future larger randomized controlled trials.

We conducted a preliminary power calculation for the given sample size based on estimates from another body awareness therapy (i.e., cognitive multisensory rehabilitation) in adults with SCI.^78^ In this prior study, we saw a reduction in neuropathic pain intensity of at least 2.31±2.07 points for the highest, average, and lowest neuropathic pain in the prior week. Assuming the same standard deviation estimate, a total sample size of 18 participants will have over 98% power to detect the same pain reduction of 2.31 points on the numeric pain rating scale (standardized Cohen’s *d*=1.12) with a two-sided significance level of 0.05 using a paired t-test, and 80% power to detect a pain reduction of 1.46 points (Cohen’s *d*=0.72) in this study. With an attrition rate of 30%, the planned sample size will still have 80% power to detect a pain reduction of 1.85 points (Cohen’s *d*=0.89).

### 2.10 Statistical analysis

Both Intent-to-Treat (ITT) and per-protocol (PP) analyses were conducted, where ITT population was defined as all participants that were enrolled in the study, and PP population was defined as all participants who were enrolled and strictly adhered to the protocol. The data analysis from this non-randomized clinical trial provides an estimate of key trial elements to determine whether to proceed to a larger definitive randomized controlled trial.

All quantitative variables are summarized using descriptive statistics at each time point. All analyses were conducted using standard statistical software including SAS, JMP 15.0, Stata, and R. The behavioral data of all participants from the first 12 weeks were analyzed using a repeated measure ANOVA, with Tukey post hoc tests to evaluate changes between every pair of time points. The data at 1Y follow-up with missingness were compared to each of the time points during the 12-week main study using the paired t-tests with Bonferroni correction of the p-values.

## 3. RESULTS

### 3.1 Feasibility measures

***Figure 1*** shows the CONSORT study flow chart of this non-randomized clinical trial. We recruited 23 adults with chronic SCI between July 1, 2021, and February 2, 2022. One participant was excluded, one declined participation, two left the study because of unrelated illnesses, and one chose not to continue. We exceeded the *a priori* set feasibility benchmarks by retaining eighteen participants (i.e., 82% retention) who completed the study, with 100% adherence to all study components, including the 6-week follow-up. Participants were reconsented to do a 1-year follow-up. This 1-year follow-up assessment was completed on February 1, 2023. Twelve out of the 18 participants agreed to participate in the 1-year follow-up assessment, six declined to participate.

**Fig. 1.**
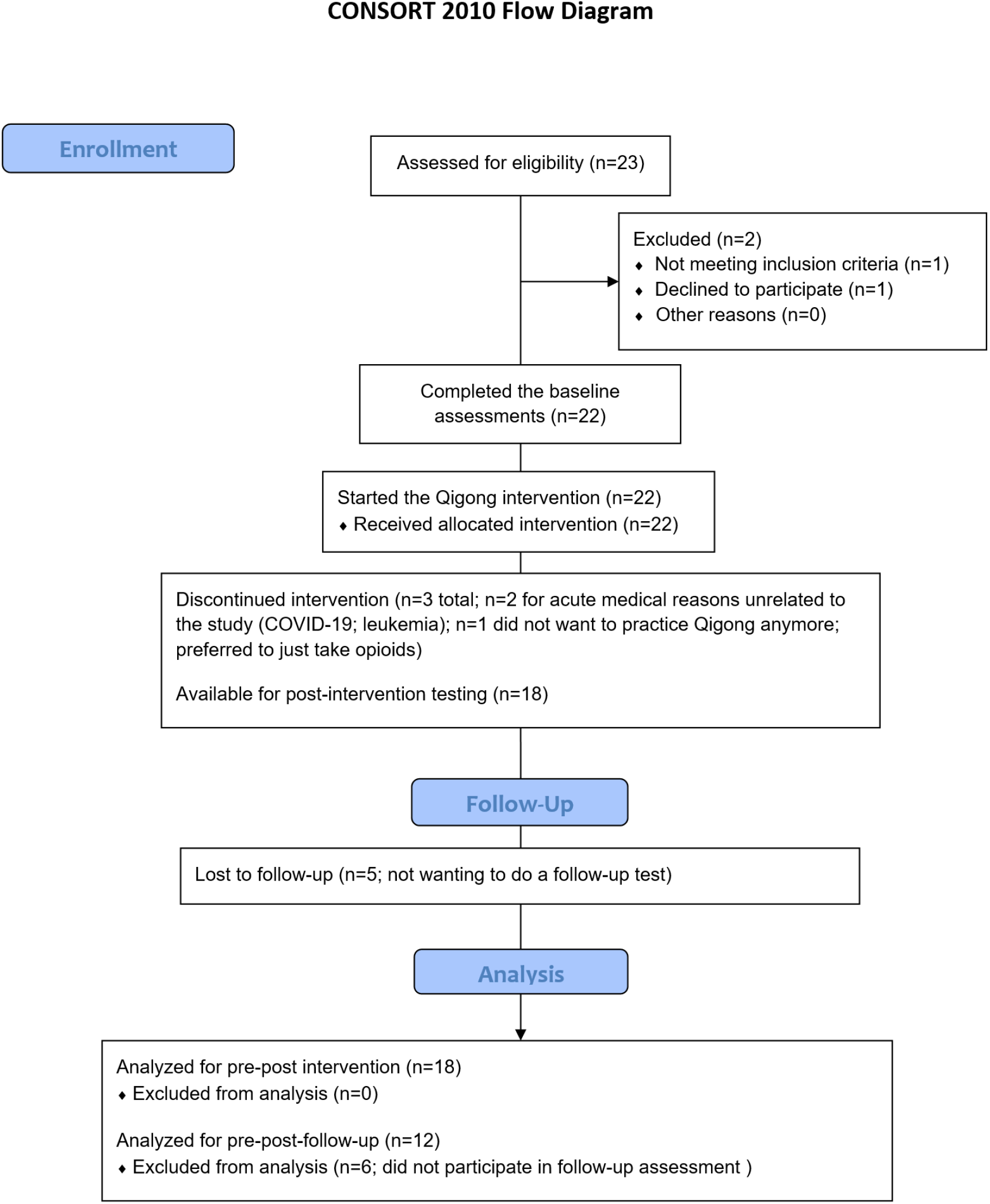
CONSORT flow diagram

Among the 18 adults, six were women, 10 with paraplegia, and 8 with tetraplegia. Thirty-three percent of adults had a complete SCI, which was slightly below our estimated 40% recruitment of adults with complete SCI. Participants had a mean age of 59.61±11.54 years (range 30-76 years of age) and were on average 15.17±11.16 years post-SCI (range 1-40 years after SCI). We enrolled a representative sample of women with SCI (33.33%), and we had a good representation of older adults, 65+ years of age (38.89%), adults living in rural areas (33.33%), and Veterans (16.67%). While we recruited participants from all over the US, all participants were non-Hispanic White, pointing to the need for more diverse recruitment strategies. About 67% had socio-economic distress and were dependent on Medicare or Medicaid. Three out of 18 (16.67%) were below the poverty threshold.^79^

Our *a-priori* benchmark of at least 70% of participants practicing at least 2x/week was exceeded: 100% of participants practiced more than 2x/week. Only two participants were just below the average of 3x/week (121.42 min and 119.85 min, respectively). All others practiced at least 123min (3x/week) with a group average of 169.72 min, or 137.98% of the required intensity of Qigong practice. There were no study-related adverse events, exceeding the *a priori* benchmark of mild adverse events in max 10% of the participants.

Participants almost unanimously reported that they could not wait to get back to the Qigong practice after the 6-week follow-up because they experienced so much benefit from it. At the 1-year follow-up, one person reported still doing Qigong every day. Five participants used Qigong when their pain increased because that was a successful tool for them to bring the pain down. Two participants developed their own methods of breathing and mindfulness, based on what they had learned in the program. Four participants had not done Qigong since the end of the intervention period.

Seventeen out of 18 participants (94.44%) were satisfied with the program, exceeding our *a priori* benchmark of 70%. One person was neutral and expressed: [a person between 56-60 years] “I feel neutral about the Qigong exercises and weekly calls. I am pain-free now and my goals (i.e., ability to perform self-defined activities, Patient Specific Functional scale) were all achieved at week 6 and were still achieved at week 12.”

### 3.2 Outcome measures

All means and standard deviations of the outcome measures are presented in ***Table 2***. The main findings are reported below. The highest baseline *neuropathic pain* was 7.94±2.33 on the NPRS^49^ (range 0-10), which was reduced to 4.17±3.07 after 12 weeks of Qigong practice (large effect size, Cohen’s *d*=1.75). Neuropathic pain was reduced by 3.78 points (large effect size, *d*=1.75), 2.06 points (large effect size, *d*=1.12), and 1.39 points (large effect size, *d*=1.08) for the highest, average, and lowest neuropathic pain in the prior week, respectively, exceeding a minimal clinically important difference for the highest and average neuropathic pain intensity levels (>1.80 points).^7,49,80,81^

**Table 1.**
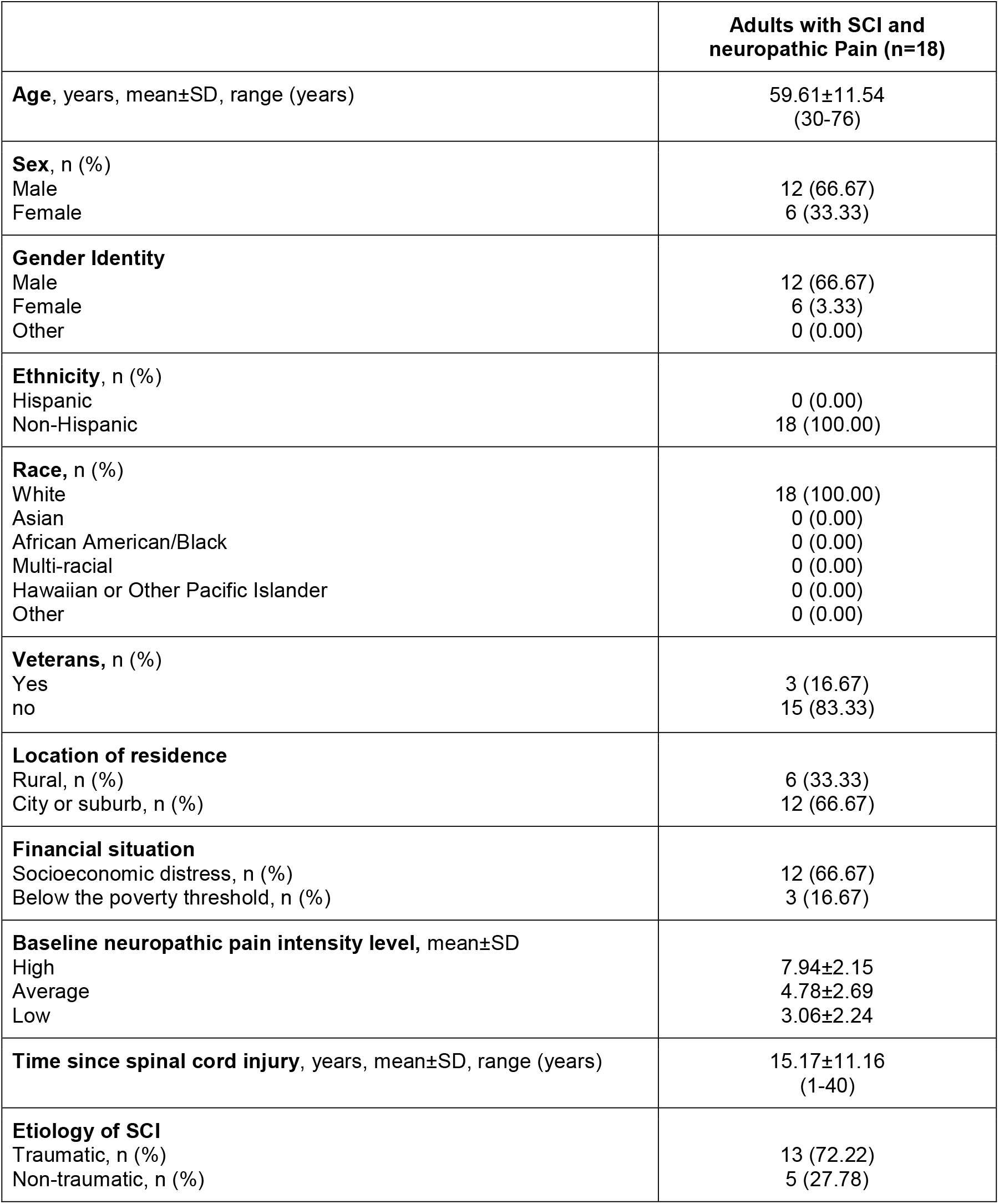
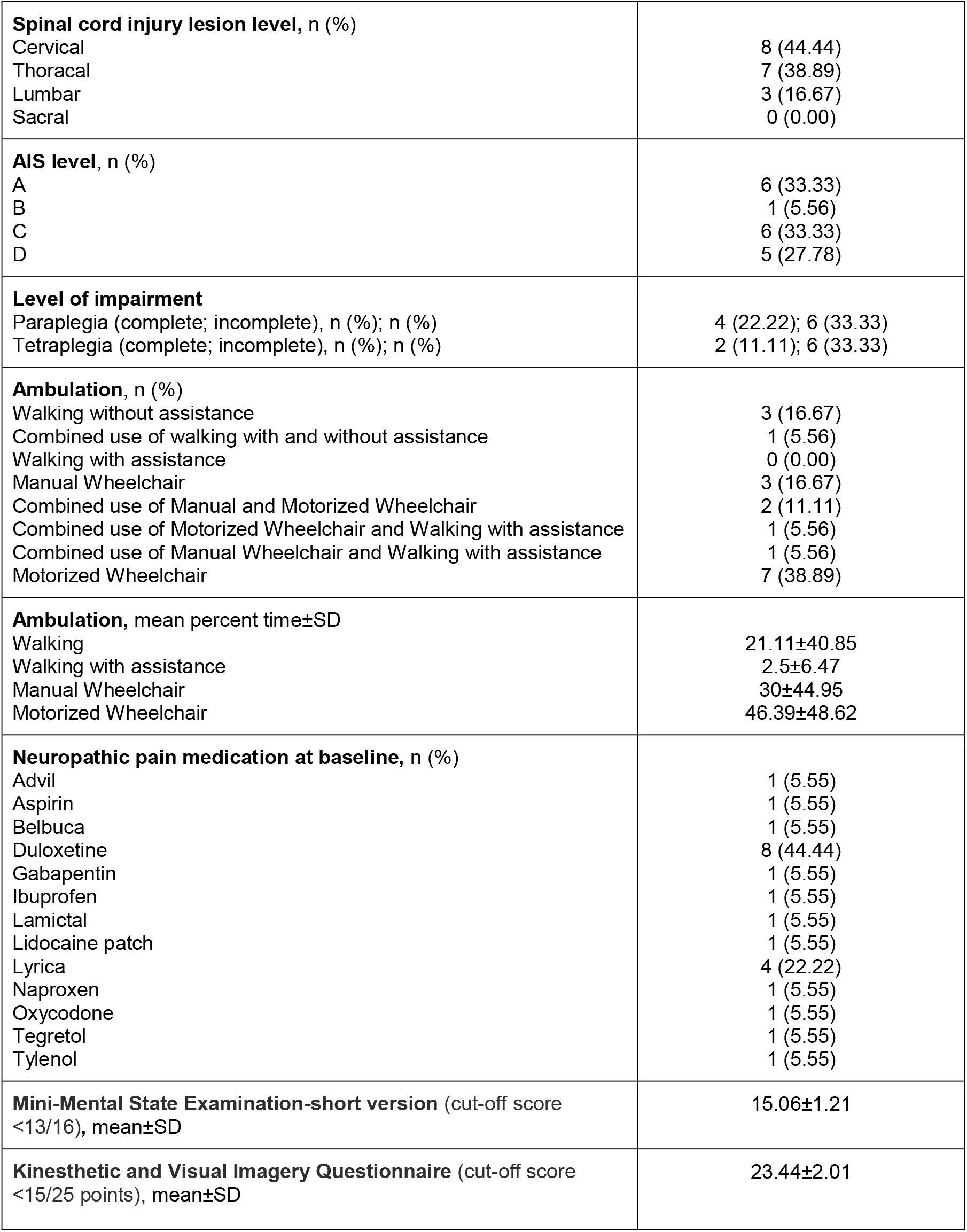
Demographic and clinical characteristics of adults with spinal cord injury and neuropathic pain.

**Table 2.** Primary and secondary outcome measures in adults with SCI-related neuropathic pain at 5 time points.

| Outcome measures | Baseline (mean±SD) | 6 weeks of Qigong (mean±SD) | 12 weeks of Qigong (mean±SD) | 6-week follow-up (mean±SD) | 1-year follow-up (mean±SD) | Repeated Measures ANOVA (all time points between baseline to 6-week follow-up) | Significant Tukey post-hoc pairs | Paired t-tests (6wFU-1YFU) |
| --- | --- | --- | --- | --- | --- | --- | --- | --- |
| n | (n=18) | (n=18) | (n=18) | (n=18) | (n=12) |  |  |  |
| Highest NPRS | 7.94±2.15 | 5.94±2.48 | 4.17±3.07 | 4.39±3.05 | 4.92±3.03 | F(3,15)=19.70<br>$p<0.0001^*$ | B-6wQ<br>B-12wQ<br>B-6wFU<br>6wQ-12wQ<br>6wQ-6wFU | t=2.24<br>$p=0.05$ |
| Average NPRS | 4.78±2.69 | 3.92±2.57 | 2.72±2.54 | 2.94±2.58 | 2.75±2.01 | F(3,15)=7.65<br>$p=0.003^*$ | B-12wQ<br>B-6wFU<br>6wQ-12wQ | t=1.74<br>$p=0.11$ |
| Lowest NPRS | 3.06±2.24 | 2.00±2.28 | 1.67±2.45 | 1.61±2.40 | 1.08±2.19 | F(3,15)=8.24<br>$p=0.002^*$ | B-6wQ<br>B-12wQ<br>B-6wFU | t=2.25<br>$p=0.81$ |
| Interference - activity | 5.39±3.55 | 3.61±2.38 | 1.94±2.29 | 2.11±2.37 | 2.67±2.84 | F(3,15)=10.09<br>$p=0.0007^*$ | B-6wQ<br>B-12wQ<br>B-6wFU<br>6wQ-12wQ<br>6wQ-6wFU | t=1.91<br>$p=0.82$ |
| - mood | 5.00±3.50 | 3.56±2.23 | 1.61±2.59 | 1.67±2.22 | 1.92±2.35 | F(3,15)=10.95<br>p=0.0005* | B-6wQ<br>B-12wQ<br>B-6wFU<br>6wQ-12wQ<br>6wQ-6wFU | t=2.24<br>p=0.05 |
| - sleep | 5.44±3.57 | 3.94±3.44 | 2.28±3.04 | 2.44±3.38 | 2.83±3.95 | F(3,15)=8.55<br>p=0.002* | B-12wQ<br>B-6wFU<br>6wQ-12wQ | t=1.43<br>p=0.18 |
| Spasm<br>- frequency | 1.67±1.37 | 1.22±1.35 | 0.50±0.71 | 0.39±0.61 | 0.50±0.90 | F(1.83,31,12)=12.71<br>p=0.0001* | B-12wQ<br>B-6wFU<br>6wQ-12wQ<br>6wQ-6wFU | t=1<br>p=0.34 |
| - severity | 1.39±1.14 | 1.06±1.16 | 0.67±0.84 | 0.33±0.49 | 0.42±0.67 | F(3,15)=7.59<br>p=0.003* | B-12wQ<br>B-6wFU<br>6wQ-12wQ | t=1<br>p=0.34 |
| PHQ-9 | 6.94±5.54 | 5.22±4.58 | 4.61±4.82 | 4.39±4.78 | NA | F(3,15)=3.48<br>p=0.04* | B-12wQ<br>B-6wFU | NA |
| PSFS | 1.07±1.46 | 4.04±2.59 | 7.76±2.85 | 7.93±2.91 | 7.64±3.19 | F(1.96,33.35)=53.18<br>p=0.0001* | B-6wQ<br>B-12wQ<br>B-6wFU<br>6wQ-12wQ<br>6wQ-6wFU | t=-0.74<br>p=0.47 |
| FAS | 3.12±0.58 | 3.13±0.60 | 3.41±0.60 | 3.52±0.51 | NA | F(3,15)=5.81<br>p=0.008* | B-12wQ<br>B-6wFU<br>6wQ-12wQ<br>6wQ-6wFU | NA |
| Autonomic<br>- general<br>function | 21.72±3.18 | 21.72±3.18 | 22.78±1.83 | 23.00±1.57 | 23.17±1.53 | $F(3,51)=5.14$<br>$p=0.004^*$ | B-6wFU<br>6wQ-6wFU | Same<br>value |
| - bladder<br>function | 1.94±2.44 | 1.94±2.44 | 2.22±2.32 | 2.22±2.32 | 2.83±2.29 | $F(3,51)=2.46$<br>$p=0.07$ | NA | $t=1.60$<br>$p=0.14$ |
| - bowel<br>function | 2.89±2.45 | 2.89±2.45 | 3.22±2.46 | 3.22±2.46 | 3.83±2.17 | $F(3,51)=2.83$<br>$p=0.05$ | NA | $t=1$<br>$p=0.34$ |
| - sexual<br>function | 2.28±2.61 | 2.28±2.61 | 2.50±2.83 | 2.61±2.85 | 3.33±3.20 | $F(3,51)=3.40$<br>$p=0.03^*$ | NA | Same<br>value |
| SCI-FI para | (n=10) | (n=10) | (n=10) | (n=10) | (n=9) |  |  |  |
| - basic<br>mobility | 26.40±9.45 | 26.80±9.81 | 31.40±5.32 | 32.80±4.05 | 31.22±6.26 | $F(1.13,10.14)=10.54$<br>$p=0.007^*$ | B-12wQ<br>B-6wFU<br>6wQ-12wQ<br>6wQ-6wFU | $t=-1.20$<br>$p=0.26$ |
| - self-care | 26.80±10.3<br>4 | 27.70±9.09 | 30.20±7.51 | 32.10±5.28 | 29.78±8.61 | $F(1.57,14.10)=8.87$<br>$p=0.005^*$ | B-12wQ<br>B-6wFU | $t=-1.07$<br>$p=0.32$ |
| - fine motor<br>function | 28.20±4.02 | 28.90±2.92 | 30.50±1.90 | 31.40±1.07 | 31.11±2.03 | $F(1.76,15.85)=7.86$<br>$p=0.005^*$ | B-12wQ<br>B-6wFU | $t=-1$<br>$p=0.35$ |
| - ambulation | 5.60±6.93 | 6.30±7.29 | 6.80±7.51 | 7.30±7.82 | 5.33±7.71 | F(1.79,16.09)=3.87<br>$p=0.05$ | NA | t=-1.79<br>$p=0.11$ |
| SCI-FI quad | (n=8) | (n=8) | (n=8) | (n=8) | (n=3) |  |  |  |
| - basic mobility | 12.00±11.2<br>6 | 13.63±12.19 | 15.00±12.8<br>2 | 15.50±12.6<br>0 | 17.67±15.04 | F(3,5)=4.03<br>$p=0.08$ | NA | Same value |
| - self-care | 14.00±13.1<br>5 | 14.25±12.71 | 17.38±13.0<br>0 | 18.00±13.0<br>2 | 22.67±10.60 | F(1.35,9.44)=7.03<br>$p=0.02^*$ | B-12wQ<br>B-6wFU | t=-2<br>$p=0.18$ |
| - fine motor function | 16.63±10.3<br>4 | 17.25±10.91 | 18.00±10.8<br>0 | 18.25±10.8<br>7 | 25.67±5.03 | F(3,5)=2.30<br>$p=0.19$ | NA | t=-1<br>$p=0.42$ |
| - ambulation | 4.00±6.30 | 4.50±6.89 | 4.63±6.93 | 4.75±7.19 | 4.33±7.51 | F(1.34,9.37)=1.63<br>$p=0.24$ | NA | Same value |

**Legend:** 12wQ: 12 weeks of Qigong practice; 6wQ: 6 weeks of Qigong practice; 6wFU: 6-week follow-up; B: Baseline; NPRS: Numeric Pain Rating Scale (used here to assess neuropathic pain intensity in the prior week); Average NPRS: pain experienced most of the time in the past week; PHQ-9: Patient Health Questionnaire-9; PSFS: Patient Specific Functional Scale; FAS: Functional Appreciation Scale; SCI/FI: Spinal Cord Injury Functional Index. Significant p-values ( $p<0.05$ ) are indicated with a \*

After the 12-week Qigong intervention, four participants were pain-free and 9 participants scored 0 on their lowest neuropathic pain. Pain reduction was maintained over the 6-week follow-up period (3 were completely pain-free, and 9 participants scored 0 on the lowest neuropathic pain intensity). The person who was pain-free after 12 weeks of Qigong but not at the 6-week follow-up reported that the increased pain was due to ongoing bladder issues and that his highest pain occurred infrequently. In total, 15 people out of 18 participants reported significant pain reduction. Those that did not reduce their neuropathic pain intensity over the course of the study shared that they enjoyed ‘listening’ or ‘watching the video’ but did not do the kinesthetic imagery or did not apply the kinesthetic imagery tools in daily life. Pressure ulcers or urinary tract infections during the study could also cause temporary increases in neuropathic pain. But even those 3 participants reported that they enjoyed watching the video and moving, that they had fewer spasms, that it was relaxing and calming, and that their sleep was better.

Repeated Measures ANOVA **(*Table 2*)** showed that there was a significant reduction in highest, average, and lowest neuropathic pain intensity levels in the prior week from baseline to post-Qigong (12 weeks of practice) and that this pain relief was maintained at the 6-week follow-up (n=18) and at 1-year follow-up (n=12). The weekly highest, average, and lowest neuropathic pain intensity ratings with the numeric pain rating scale (NPRS)^49^ are shown in ***Figure 2***. Some participants had urinary tract infections or had surgeries at 1-year follow-up, causing a temporary increase in highest neuropathic pain levels.

**Fig. 2.**
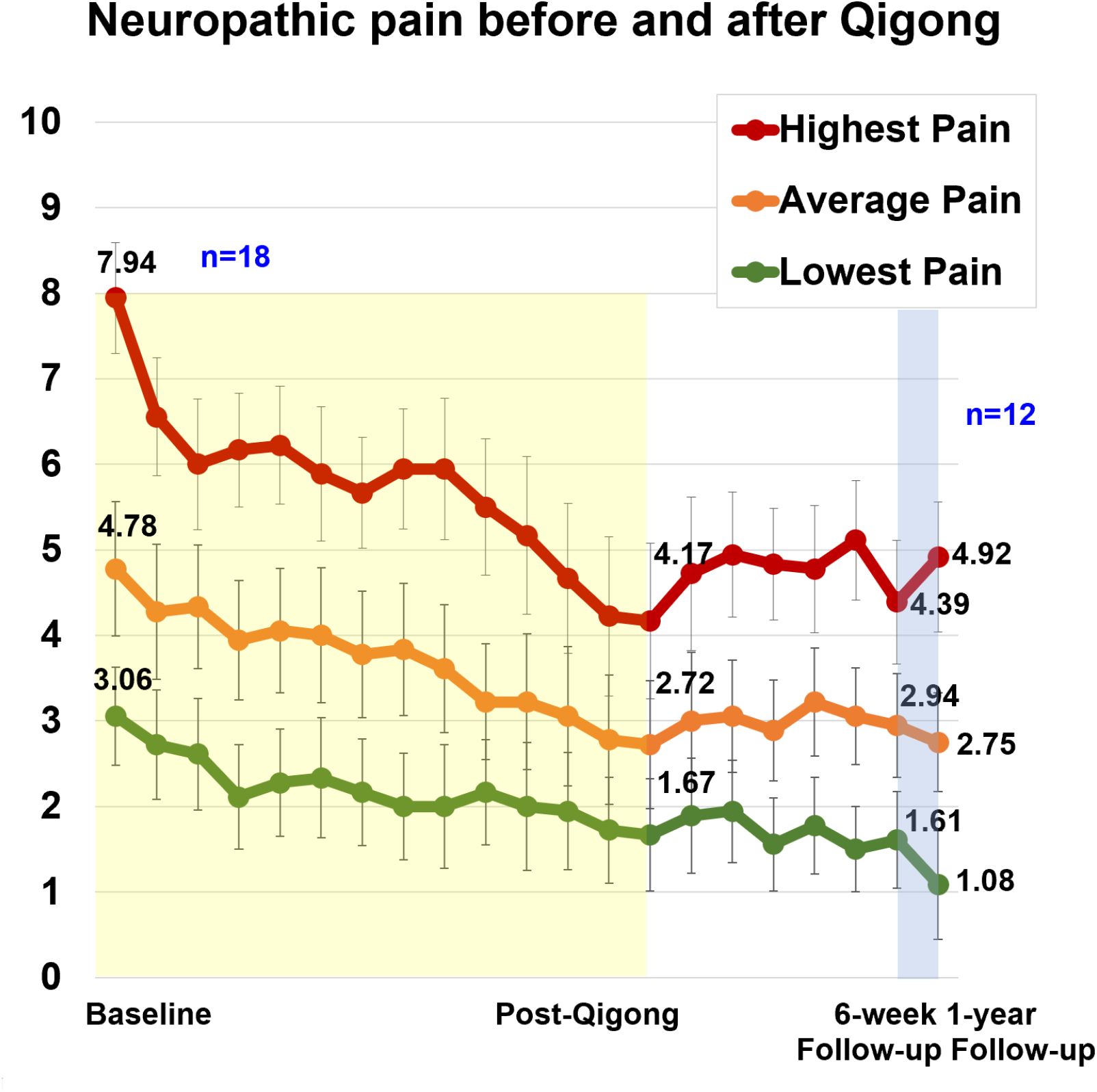
Weekly neuropathic pain rating levels for highest, average, and lowest pain in 18 adults with SCI. The 12-week Qigong training is highlighted in yellow. During the 6-week follow-up (n=18) participants did not practice Qigong but implemented body awareness in daily life. Afterwards they could resume Qigong practice at the frequency of their choice. Their neuropathic pain levels at 1-year follow-up (n=12) are shown in the line graph as well.

Aside from pain reduction, participants reported reduced spasm frequency (change score 1.17±1.20, large effect size, *d*=0.98) and severity (change score 0.72±1.02, moderate effect size, *d*=0.71), and reduced interference of neuropathic pain on activity (change score 3.44±2.53, large effect size, *d*=1.36), mood (change score 3.39±2.40, large effect size, *d*=1.41), and sleep (change score 3.17±2.77, large effect size, *d*=1.14), measured by the NINDS-CDE International SCI Pain Basic Data Set Version 2.0 (***Figure 3)***.^66^

**Fig. 3.**
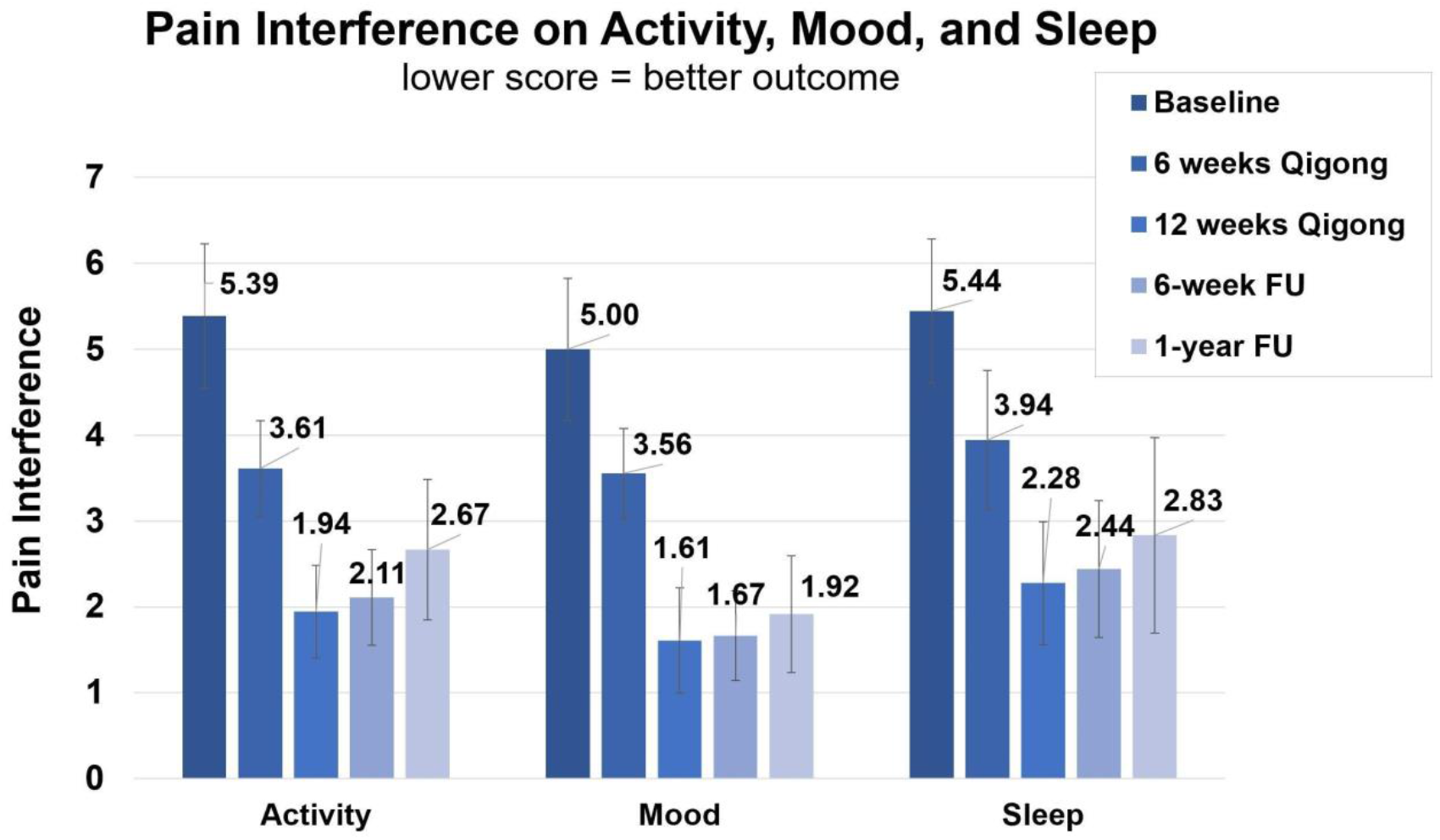
Clinical assessments of the NINDS-CDE International SCI Pain Basic Data Set Version 2.0.1. Average score of interference of neuropathic pain with activity, mood, and sleep. Assessments were taken at baseline, 6-, and 12-weeks of Qigong practice, 6-week-, and 1-year follow-up.

Participants performed better on functional activities that were important to them (PSFS, change score 6.68±3.07, large effect size *d*=2.18, ***Figure 4***) and had improved mood (PHQ-9^72^, change score 2.33±3.31, moderate effect size, *d*=0.70). Participants also improved on the functional appreciation scale (change score -0.29±0.48, moderate effect size, *d*=0.60).

**Fig. 4.**
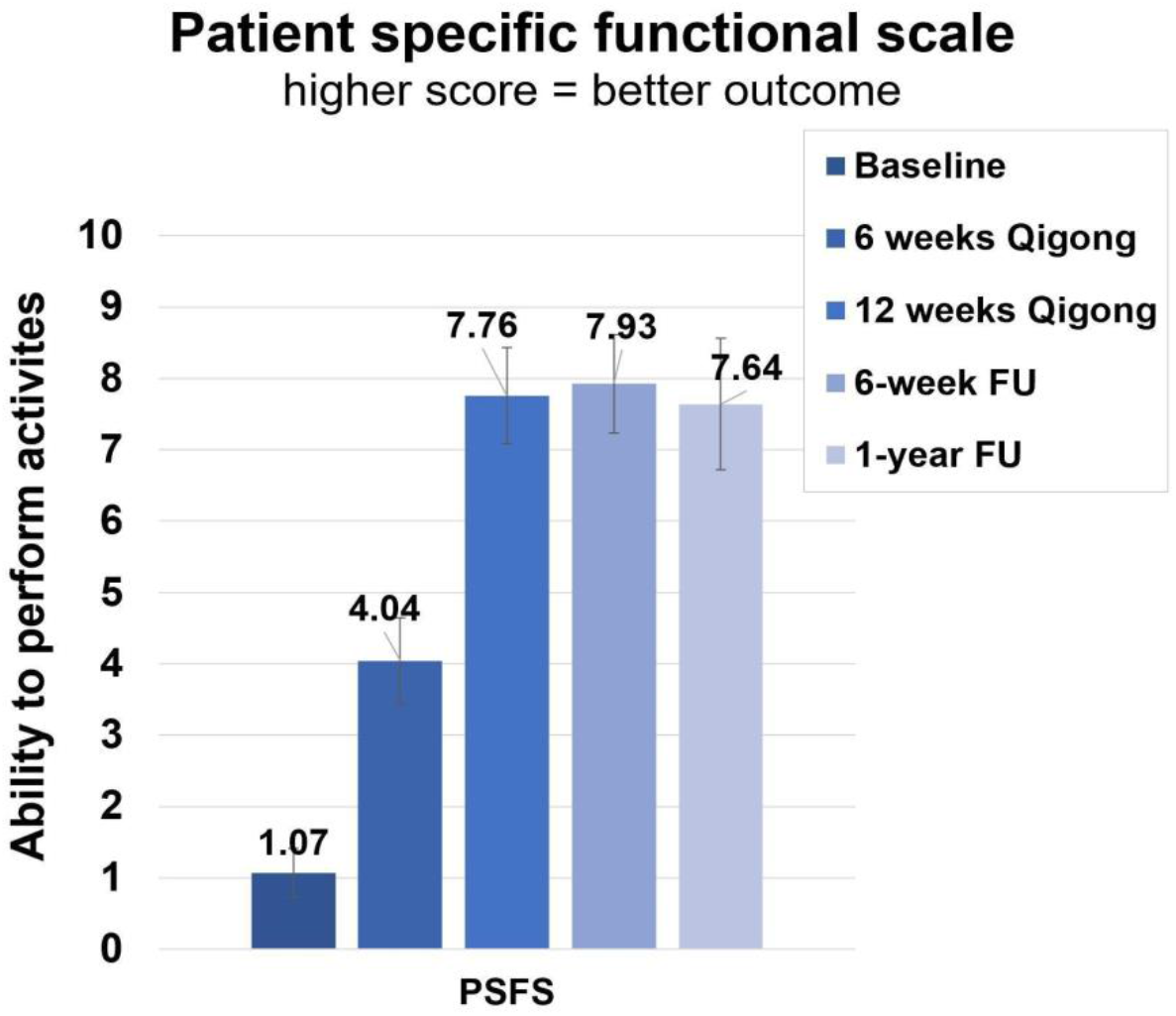
Clinical assessments of the Patient Specific Functional Scale (PSFS). Average score of ability to do three functional activities. Assessments were taken at baseline, 6-, and 12-weeks of Qigong practice, 6-week-, and 1-year follow-up.

An example of a functional activity, where one participant reported improvements, was being able to spend at least 6 hours and even overnight with the grandkids (small children). At baseline, this participant could only spend 2-3 hours with their grandkids before she had neuropathic pain. After 12 weeks of Qigong practice, she spent a whole weekend with 3 grandkids including a sleep-over. Another person’s goal was to be able to exercise for 30 minutes without neuropathic pain. At baseline, the participant always felt neuropathic pain during exercise, causing him to fatigue easily. After 12 weeks of Qigong practice, he was able to exercise for 30 minutes without neuropathic pain.

Overall, the participants reported more intense daily activity (e.g., reorganizing the furniture in the house and cleaning; relocating and organizing a move with children; helping their children move). Family members also benefited. One mother reported that her two young children were doing the Qigong practice with her. Another person could play the piano again after 3 years, his favorite hobby. He bought a new adjustable piano bench and created a slant so that his pelvis was oriented forward helping with better positioning and, thanks to the Qigong practice, he was able to feel his feet. He reported that he now plays more than 30 minutes/day. This participant practiced Qigong every day and was pain-free after 12 weeks. He did not have spasms anymore, slept better, was calmer, and could handle things better.

The results from the post-hoc tests after the Repeated Measures ANOVA analysis for all time points from baseline to 6-week follow-up (n=18) showed that general cardiovascular function improved between baseline and 6 weeks follow-up (n=18) and those results were maintained at 1-year follow-up (analyzed by paired t-tests between 6-week and 1-year follow-up). Empirical reports of improvements in general functioning had to do with temperature regulation. Four participants reported improved bowel function through the course of the study, which had to do with the fact that they could feel better when there was a need for bowel movements (greater interoception), improved sphincter control and they were better at avoiding leakage. Four other participants reported they could feel more (and enjoy more) during sexual activity. These reports were backed up by the data showing a significant improvement over time for sexual function, analyzed with repeated measures ANOVA. Two participants reported increased awareness of the need to urinate, or feeling urine passing through the catheter. Several participants reported that after the Qigong intervention, they became more acutely aware when something was ‘off’ in their bodies and thus they could address urinary tract infections, or uncomfortable seating positions quicker.

Finally, participants with paraplegia reported improved balance, which was reflected in improved scores in basic mobility, self-care, and fine motor function on the SCI/FI assessment. An example of improved fine motor function was that they did not need to use their hands anymore for balance and thus could do fine motor activities while sitting in the wheelchair. Participants with tetraplegia improved in self-care.

## 4. Discussion

The results of this non-randomized controlled trial demonstrate the feasibility of the design and implementation of a remote Qigong intervention, and adherence to a 12-week intervention with a 1-year follow-up in 18 adults with SCI-related neuropathic pain. The 12 weeks were well tolerated, there were no study-related adverse events. The vast majority of participants enjoyed the Qigong practice and continued to use it as needed in daily life afterward, pointing to a change in behavior that helped them gain control over an otherwise debilitating symptom. Below is feedback from participants who enjoyed the Spring Forest Qigong™ practice:

[a person between 56-60 years], “It got me back in touch with meditating that I felt I was missing in my life. And then at times I have more body awareness which can further increase over time. I have much less pain. I am grateful and glad that I was able to do it.”

[a person between 66-70 years] *“*The fact that the pain is reduced makes life more pleasurable, especially with social contacts (family gatherings).*”*

[a person between 51-55 years] *“*Qigong is relaxing and calming and that relaxed and calm feeling carries over into the day.*”*

[a person between 75-80 years] *“*Two thumbs up. I can’t say enough good about it, how many good things it is doing for me. I am practicing almost every day. Before the study I was ready to jump from a building (‘ready to get out of this body’) and now I do not have these feelings anymore. The underlying results are excellent: pain-free, no spasms, sleep is good now, calmer, I can handle things better. I can play the piano again after 3 years. Before the study I used to be antsy after about 15 min at the computer and I had to take a nap on the bed. Now I can sit at the computer for 3-4h and it feels good. Also at night, I used to be cold and needed a sweatshirt to sleep. Now I stay comfortably warm at night and I do not need the sweatshirt anymore. In the last 2 weeks, I also regained sensation of the catheter. I feel when urine is passing through which I could not feel before.*”*

[a person between 45-50 years] *“*It was really helpful to get more in tune with my body. This helped me figure out that something was not right and to get iron infusions that have helped. My pain is down and I am sleeping better. I knew that if I signed up for the study I would do it, because of outside accountability. There was curiosity involved. The Qigong was relaxing and soothing. It was helpful for my children as well. The plantar fasciitis is better. It is healing faster. The tingling in the feet is better and the pain is down so I am sleeping better. I am more social, I have more energy, and organize things easier. I have more control over life. It is easier now for me to make decisions and to undertake a big thing like a move.*”*

[a person between 55-60 years] *“*I am over the moon! I never thought that it would go this well. Before the study, I was spending much time in the shower to deal with the pain and I have had no need at all anymore for that. My feeling has been that it was absolutely wonderful! I am so happy for the opportunity to be part of this study because it made so much difference. I do not need to lay in the shower anymore to reduce the pain and I was able to sit for a whole day, inspiring students and researchers by sharing my journey. I told them: And here is this study… what a difference it is making in people’s lives. The idea that there is something out there that could help me, considering how long ago I had the SCI (29 years of dealing with this pain!)… and here in only 8 weeks it makes a profound difference in my life. It gives people hope. Prior to the study, I would not have been able to sit here and talk to you for so long. This is another proof of how profound the difference is that this study has made.*”*

[a person between 55-60 years] *“*What you practice grows stronger. It stayed with me. The positivity part has been more prominent there for me. My life is changing a lot. I have been able to realize that those 5 emotions from the 5 Element Healing Movements are what I want for my life. There are changes in friendship. I have stopped taking calls from negative people and spent more time with positive people. As simple as those points are, they are profound. Years ago, a therapist told me that you can decide in the morning how you want to use your energy. That makes sense now. I have been using energy now to bring joy and contentment. I have been a lot more active, I do not have to lay down anymore during the day.*”*

Twelve weeks appears to be the best duration for sustained neuropathic pain reduction based on post hoc testing (***Table 2***), which shows a significant reduction of highest and lowest neuropathic pain at 6 weeks, but the reduction of average pain in a week (i.e., pain experienced most of the time) becomes significant at 12 weeks of Qigong practice. Given that our group exceeded, on average, the requested time to practice (3x/week), this frequency of practice is feasible for the majority of participants.

It seems that the adaptation of the Qigong practice for adults with SCI by combining active movements to their ability level with kinesthetic imagery (i.e., focusing on the feeling in the body as if doing the whole-body Qigong movement while standing) seemed particularly effective and worth pursuing further in a larger study.

Aside from pain reduction and improvements in mood and function, participants also reported a change in mindset in how they approached their pain. Before the Qigong study, participants tended to distract themselves from the pain or ignore the pain, and, as a result, often the pain worsened. After the Qigong practice, many participants shared that they learned how to connect with their bodies, developed the ability to listen to their bodies, and understand when their bodies gave signals of an uncomfortable position or situation (e.g., upcoming bowel or bladder issue). Participants felt they reacted quicker to these signals and the uncomfortable feeling did not develop into pain.

There are some limitations to this study. This non-randomized clinical trial was conducted in a small sample and thus further validation in a larger randomized trial is needed. Furthermore, while we were able to recruit participants who live in remote areas and/or experience financial distress, diversity in terms of race and ethnicity was missing. For example, adults of Hispanic background comprise 17.4% of the US population (55.4 million)^82^ and represent 8.3% of all SCI since 2005;^83,84^ yet, they are underrepresented in SCI rehabilitation studies.^1^

## Conclusions and Relevance

Our preliminary data demonstrate the feasibility and acceptability of practicing Qigong for adults with SCI-related neuropathic pain, generating promising results in terms of neuropathic pain reduction and improvements in SCI-related symptoms. The data from the present work will inform the design of future randomized controlled trials with adequate sample size, intervention dosage, and follow-up duration.

The remote delivery of Qigong offers multiple applications for broad use in the home or community. Further studies in adults with SCI of different races and ethnicity, as well as in other languages (such as Spanish) are needed.

## Data Availability

All data produced in the present study are available upon reasonable request to the authors.

## Trial Registration (this manuscript refers to the quasi-experimental substudy)

CREATION: A Clinical Trial of Qigong for Neuropathic Pain Relief in Adults with Spinal Cord Injury, NCT04917107, https://www.clinicaltrials.gov/ct2/show/NCT04917107.

## Acknowledgments

Our gratitude goes to Qigong Grand Master Chunyi Lin, MS, and Qigong Master Jaci Gran from the Spring Forest Qigong™ for their consultancy and assistance with providing the teaching and content of the “5 Element Healing Movements”. We thank Debra Lin and Collin Silas for their administrative and logical assistance. We thank our participants and all adults with SCI-related neuropathic pain who gave their valuable input on the research questions, intervention, assessments, and elements of the study design. We present our profound thanks to Marc Noël for the critical review of the manuscript.

